# The risk of COVID hospital admission and COVID mortality during the first COVID 19 wave with a special emphasis on Ethnic Minorities: an observational study of a single, deprived, multi ethnic UK health economy

**DOI:** 10.1101/2020.11.20.20224691

**Authors:** Baldev M Singh, James Bateman, Ananth Viswanath, Vijay Klaire, Sultan Mahmud, Alan M Nevill, Simon J Dunmore

## Abstract

**Objectives:** To address the generalisability of COVID-19’s outcomes to the well-defined but diverse communities of a single City area.

**Design:** An observational study of COVID-19 outcomes using quality-assured and integrated data from a single UK hospital contextualised to its feeder population and its associated factors (comorbidities, ethnicity, age, deprivation).

**Setting/Participants:** Single city hospital with a feeder population of 228,632 adults in Wolverhampton’s city area.

**Main Outcome Measures:** Hospital admissions and mortality.

**Results:** 5558 patients admitted, 686 died (556 in hospital); 930 were COVID-19 admissions (CA),of which 270 were hospital COVID deaths, 47 non-COVID deaths, 36 deaths post-discharge; 4628 non-COVID-19 admissions (NCA), 239 in-hospital deaths (2 COVID), 94 deaths post-discharge. 223,074 adults not admitted, 407 died. Age, gender, multi-morbidity and Black ethnicity (OR 2.1 [95% CI 1.5-3.2] p<0.001, absolute excess risk of <1/1,000) were associated with COVID-19 admission and mortality. The South Asian cohort had lower CA and NCA, lower mortality (CA (0.5 [0.3-0.8], p<0.01), NCA (0.4 [0.3-0.6] p<0.001), community deaths (0.5 [0.3-0.7] p<0.001). Despite many common risk factors for CA and NCA, ethnic groups had different admission rates, and within-groups differing association of risk factors. Deprivation impacted only in White ethnicity, in the oldest age bracket and in a lesser (not most) deprived quintile.

**Conclusions:** Wolverhampton’s results, reflecting high ethnic diversity and deprivation, are similar to other studies for Black ethnicity, age and comorbidity risk in COVID-19 but strikingly different in South Asians and for deprivation. Sequentially considering population and then hospital based NCA and CA outcomes, we present a complete single health-economy picture. Risk factors may differ within ethnic groups; our data may be more representative of communities with high BAME populations, highlighting the need for locally focussed public health strategies. We emphasise the need for a more comprehensible and nuanced conveyance of risk.

**Strengths and limitations of this study:**

- The rapidly developing COVID-19 pandemic has led to numerous studies (published, preprints and national public health reports) of its health impacts in relation to ethnicity, co-morbidities and other factors; few studies, however, have attempted to evaluate infection patient data in terms of morbidity and mortality in context of the feeder population and most are limited by incompleteness of data and inability to account for regional variations in factors such as ethnicity and deprivation
- Our observational study used a high quality and complete dataset from the local population and the hospital serving it to examine the association of purported risk factors with severity and mortality and the results reveal the importance of evaluating such risks in the local, and not just national, population setting taking into account the local variations in patient backgrounds
- We found an increased risk of COVID-19 mortality for Black ethnicity (OR 2.1) but a decreased risk (OR 0.5) for South Asians, compared with white ethnicity; Our analysis reveals that a nuanced approach to studying risk factors associated with COVID-19 severity and mortality is important – factoring in regional variation in ethnicity, deprivation etc. specifically linked to the source population
- We suggest, based on our findings, that understandably rapid analysis and dissemination of studies of COVID-19 risk needs to be tempered by careful consideration of the real implications; we further urge caution in conveying risk messages to the wider community because of an ethical imperative to ensure such messages do not lead to unnecessary fear and deter individuals, particularly from specific ethnic backgrounds, from seeking needed medical assistance.

## Introduction

In understanding the natural history of disease, fundamental to healthcare, the COVID-19 (hereafter referred to as “COVID”) pandemic highlights issues within data repositories. Constructing multiple source datasets has complexity in case definition, data acquisition, integration, quality, completeness, coding accuracy and the clinical meaning of analysis outcomes. ^1-4^ Emphasising this challenge, national UK data were initially collated via the Patient Notification System, requiring a positive swab test up until the 28^th^ April 2020 but revised to include clinical definitions given an estimated false negative rate testing rate of up to 29%. ^5-8^ Well-established primary care databases, may have significant inaccuracy and do not include hospital secondary care information.^9^ A large UK primary care epidemiological study also used national COVID (SARS-CoV-2) positive swab cases for case definition.^2^ Conversely, secondary care case series and international registry studies for specific diseases are not linked to primary care datasets.^10-14^ Therefore important caveats exist in utilising and interpreting such data and drawing clinically important conclusions regarding the adverse associations of ethnicity with outcomes.^2, 15^

Our objective, therefore, was to establish a tightly governed comprehensive, multi-source, integrated, quality assured local structured clinical data set, used for the purposes of direct care, define cohorts at risk, to systematically improve clinical coding and mortality recording accuracy, and to enable an informed understanding of factors influencing hospital activity, including admissions. This approach should ultimately inform public health initiatives. We present a proof of principle study to evaluate the utility of this approach in relation to a single UK city wide health district, reporting our findings regarding population wide factors that may have an association with 2 key COVID outcomes, hospital admission and mortality, over the first 12 weeks of the pandemic in this City.

## Methods

### General Method

The time frame spanned 1/3/2020 to 24/05/2020.

Data were integrated into an SQL database from primary care, community and hospital clinical and pathology systems for all people resident in Wolverhampton or registered to Wolverhampton practices and those from immediately adjacent districts with emergency admission to New Cross Hospital (NXH). Only those alive at the start point were included and subsequently death and date of death were tracked. The final total population aged >18 was 228,632, of whom 1063 were resident but not Wolverhampton GP registered, 1521 who were registered but not City resident and 1026 neither resident nor registered from immediately surrounding areas with an emergency admission to NXH, such that 99.5% of the cohort were registered and/or resident constituting 88% of COVID admissions and 91% COVID deaths. The Index of Multiple Deprivation was allocated according to postcode. Unavailable smoking status (15%) was reallocated to “non-smoker”. Missing body mass index (22%) were replaced by the age-related (5-year band) mean value in the cohort. Ethnicity data from all sources were reviewed, only unambiguous data were accepted, and recoded into Caucasian (White), South-Asian, Black, Mixed Ethnicity Chinese with 7.5% remaining “Unknown”. Comorbidities were accrued and cross-checked from primary care and hospital coding to include: Asthma, COPD, Diabetes, Hypertension, Coronary Artery Disease, Stroke and Peripheral Arterial Disease, Chronic Heart Failure, Atrial Fibrillation, Chronic Kidney Disease, Cancer, Dementia, Depression, other Mental Health Disorders, Epilepsy, Learning Difficulties, Osteoarthritis, Rheumatoid arthritis as well as recorded nursing home residency and palliative care status. Non-elective admissions over the preceding 12 months were ascertained. During admission, the COVID clinical status was recorded by the Infection Diseases team or the clinical team in daily updates as “COVID definite”, “COVID probable” or “not COVID”. Formal endpoint coding was in duplicate with a rolling triangulation audit in place comparing the clinical diagnosis, the coded diagnosis and COVID pathology status for coding accuracy. Mortality and cause of death were certified in our Medical Examiner System and also continuously cross-checked against the coded status. COVID coding and death certification arbitration were supported by the accountable senior responsible consultant (AV). Further validation against the National Strategic Tracing Service captured deaths outside hospital.

### Statistical Analysis

This was undertaken in SPSS v26. Factors analysis of all variables considered confounding effects and redundancy, yielding a 9 component rotated solution explaining 48% of the variance: deprivation and ethnicity were strongly co-associated in a single component whilst the two principal outcome measures of hospital admission and mortality were in another distinct component. We adopted a multinomial regression analysis approach. The analysis was undertaken sequentially to ensure an a *priori justification* for further analysis. Statistical tests are described in the text and their results considered significant at p<0.05.

### Ethical Approval

This was not sought nor deemed necessary since this work represents a continuous quality improvement programme of the informatics component of service changes required between various local NHS organisations for integrated working stipulated during the COVID 19 emergency. Data governance was in line Trust policy and with the COVID emergency directive of NHS England.

### Patient and Public Involvement

None (not applicable to this type of study)

## Results

### Hospital Admissions

The population characteristics are shown in Table 1, grouped according to admission status (No Admission, Non-COVID Admission and COVID Admission (NA, NCA, CA respectively) together with their mortality rates. Compared to NA, there was an increased association of all variables from NCA through to CA including age, the number of comorbidities, most individual comorbidities, the surrogate measures of dependency, and prior history of emergency admissions. Male gender, BMI, IMD and smoking status were significantly different. Ethnic minority groupings were significantly different between admission types with the South Asian population prevalence in CA being 46% of that in the comparator NA population whilst the Black population appeared to have a 56% excess. Table 2 gives further numerical detail.

**Table 1.** The demographic, clinical features and mortality outcomes of a whole adult population (n= 228,632) categorised according to their hospital admission status during 12 weeks of the UK COVID-19 pandemic. Data are presented as the mean+-SD or as percentages. Between groups analysis is by ANOVA or by Chi square for scale or categorical variables respectively. Co-morbidities are listed in descending order of frequency.

| Table 1 | Whole population | Not admitted | Non COVID Admission | COVID Admission |  |
| --- | --- | --- | --- | --- | --- |
| Number (% of total) | 228,632 | 223,074 (97.6%) | 4,628 (2%) | 930 (0.4%) |  |
| Age (years) | 50.0 ± 18.8 | 47.6 ± 18.5 | 63.1 ± 20.5 | 71.4 ± 16.5 | p<0.001 |
| Gender (male, %) | 114,866 (50.2%) | 50.3% | 48.4% | 55.2% | p<0.001 |
| Ethnicity (% in category) |  |  |  |  |  |
| · White | 142,781 (62.5%) | 62.1% | 77.7% | 73.5% | p<0.001 |
| · South Asian | 44,229 (19.3%) | 19.6% | 10.4% | 8.9% |  |
| · Black | 17,858 (7.8) | 7.9% | 4.6% | 12.2% |  |
| · Mixed | 5,809 (2.5%) | 2.6% | 1.3% | 0.6% |  |
| · Chinese | 806 (0.4%) | 0.4% | 0.1% | 0.2% |  |
| · Unknown | 17,149 (7.5%) | 7.5% | 5.8% | 4.5% |  |
| Index of Multiple Deprivation | 32.8 ± 15.9 | 32.9 ± 15.9 | 30.4 ± 14.4 | 30.7 ± 14.4 | p<0.001 |
| Smoking status (% never) | 148,046 (64.8%) | 65% | 73% | 81% | p<0.001 |
| Body Mass Index (kg/m<sup>2</sup>) | 27.3 (± 5.7) | 27.3 (± 5.7) | 27.8 (± 5.1) | 27.9 (± 4.6) | p<0.001 |
| Prior admission (Any 1 year), (≥3) (%) | 15,119 (7%) (0.6%) | 6% (0.4%) | 31% (7%) | 35% (8%) | p<0.001 |
| Any comorbidity (≥3) (%) | 110,564 (48 %) (12 %) | 48% (11%) | 77% (41%) | 88% (58%) | p<0.001 |
| Palliative care registered | 1,530 (0.7%) | 0.5% | 4.9% | 13.9% | p<0.001 |
| Nursing home resident | 2,130 (0.9%) | 0.8% | 4.7% | 9.7% | p<0.001 |
| Hypertension | 47,830 (20.9%) | 20.2% | 47.6% | 64.2% | p<0.001 |
| Depression | 39,153 (17%) | 17.0% | 22.4% | 18.7% | p<0.001 |
| Asthma | 30,335 (13.3%) | 13.2% | 16.1% | 16.3% | p<0.001 |
| Diabetes | 20,529 (9.0%) | 8.6% | 21.4% | 33.8% | p<0.001 |
| Ischaemic heart disease | 10,965 (4.8%) | 4.4% | 19.9% | 23.2% | p<0.001 |
| Chronic kidney disease | 10,265 (4.5%) | 4.1% | 17.6% | 29.0% | p<0.001 |
| Cancer | 9,796 (4.3%) | 4.0% | 16.0% | 17.8% | p<0.001 |
| Atrial fibrillation | 6,771 (3.0%) | 2.6% | 4.7% | 8.7% | p<0.001 |
| Chronic obstructive pulmonary disease | 6,762 (3.0%) | 2.7% | 12.3% | 15.1% | p<0.001 |
| Cerebro-vascular disease | 5,359 (2.3%) | 2.2% | 8.8% | 13.5% | p<0.001 |
| Cardiac failure | 4,940 (2.2%) | 1.9% | 13.1% | 20.8% | p<0.001 |
| Osteoarthritis | 4,267 (1.9%) | 1.7% | 8.0% | 11.2% | p<0.001 |
| Epilepsy | 4,035 (1.8%) | 1.7% | 3.8% | 4.5% | p<0.001 |
| Rheumatoid arthritis | 3,284 (1.4%) | 1.4% | 4.2% | 6.2% | p<0.001 |
| Mental health disorder | 2,967 (1.3%) | 1.3% | 1.7% | 1.7% | p<0.05 |
| Peripheral vascular disease | 2,724 (1.2%) | 1.1% | 4.7% | 8.7% | p<0.001 |
| Dementia | 2,304 (1.0%) | 0.9% | 4.0% | 7.8% | p<0.001 |
| Learning difficulties | 1,597 (0.7%) | 0.7% | 1.0% | 1.2% | p<0.05 |
| Vital status (n, % died) | 1,093 (0.5%) | 407, 0.2% | 333, 7.2%<sup>s</sup> | 353, 38%<sup>+</sup> | p<0.001 |

<sup>s</sup>, 237 non-COVID hospital deaths, 94 post discharge deaths in community, 2 hospital COVID deaths; <sup>+</sup>, 270 hospital COVID deaths, 47 non-COVID hospital deaths, 36 post discharge deaths in community

**Table 2.** COVID admission and death by ethnic category showing numbers, absolute rates per 1000 population, and the excess risk and unadjusted OR s (95% CI) vs the White group as comparator.

| Table 2 | White | South Asian | Black | Other or Unknown |
| --- | --- | --- | --- | --- |
| Total numbers | 142,781 (63%) | 44,229 (19%) | 17,858 (8%) | 23,764 (10%) |
| COVID admission | 684 (74%) | 83 (9%) | 113 (12%) | 50 (5%) |
| COVID admission/ 1000 | 4.8 | 1.9 | 6.3 | 2.1 |
| COVID admission excess risk / 1000 | comparator | -2.9 | 1.5 | -2.7 |
| COVID admission OR | comparator | 0.39 (0.31 - 0.48), p<0.001 | 1.31 (1.07 - 1.59), p<0.001 | 0.43 (0.33 - 0.58), p<0.001 |
| COVID death | 190 (70%) | 32 (12%) | 39 (14%) | 11 (4%) |
| COVID death/ 1000 | 1.3 | 0.7 | 2.2 | 0.5 |
| COVID death excess risk / 1000 | comparator | -0.6 | 0.9 | -0.9 |
| COVID death OR | comparator | 0.54 (0.37 - 0.79), p<0.01 | 1.64 (1.16 - 2.31), p<0.01 | 0.35 (0.19 - 0.64), p<0.01 |
| COVID death / COVID admission / 1000 | 0.3 | 0.4 | 0.3 | 0.2 |
| COVID death in COVID admission OR | comparator | 1.55 (0.96 - 2.51), p=0.075, ns | 1.19 (0.78 - 1.83), p = 0.423, ns | 0.63 (0.32 - 1.25), p=0.187, ns |

The 3 hospital admissions categories (NA, NCA, CA) were taken as the response variable and submitted to multinomial regression (Table 3). The complete model was highly significant (χ^2^=8,869.1, p<0.001). Male gender was more prevalent in CA. Age distribution (Figure 1a) differed significantly for CA and NCA versus NA, and the two admission groups differed significantly from each other, reflecting the higher mean age in CA. The pattern for deprivation (Figure 1b) showed the peak admission rates to be in the second least deprived quintile with the most deprived quintile not being significantly different from the least deprived quintile, whilst the 2 admission groups did not differ significantly from each other in this regard. There was a decreased relative risk for admission in either group with current or previous smoking. Both admission groupings had a significantly increased history of prior emergency admissions, established multi-morbidity, being nursing home resident or in a palliative phase of care with these latter two characteristics in significantly higher prevalence in CA compared to NCA. Both groups shared individual comorbidities in higher risk, but with some differential effect for diabetes, hypertension, atrial fibrillation and peripheral vascular disease which were increased in CA.

**Table 3.** Multinomial regression for the association of factors with COVID or Non COVID related emergency hospital admissions (HA) compared to the reference category of those not admitted (n=223, 074). Data are the Odds Ratio (OR) with 95% confidence intervals. For age and IMD as categorical ordinal variables (data ranges shown), the comparators were the youngest and least deprived quintiles respectively. Comorbidities associations are listed in descending OR order for the CA group. The comparison of CA vs NCA was by binary logistic regression. Variables not listed (Table 1) were excluded stepwise as not significant.

| Table 3 | COVID HA | Non COVID HA | CHA vs NCHA |
| --- | --- | --- | --- |
| Number (%of population) | 930 (0.4%) | 4,628 (2%) |  |
| Gender (male) | 1.5 (1.3 - 1.7), p<0.001 | ns, p=0.48 | 1.3 (1.1 - 1.5), p<0.001 |
| Age category Q2 (30-40) | 2.7 (1.5 - 5), p<0.01 | 1.2 (1 - 1.3), p<0.05 | 2.2 (1.2 - 4), p<0.05 |
| Age category Q3 (41- 51) | 5.1 (2.9 - 9), p<0.001 | 1.4 (1.2 - 1.6), p<0.001 | 3.2 (1.8 - 5.8), p<0.001 |
| Age category Q4 (52 - 65) | 7.8 (4.6 - 13.4), p<0.001 | 1.7 (1.5 - 1.9), p<0.001 | 3.7 (2.1 - 6.5), p<0.001 |
| Age category Q5 (66 - 113) | 16.6 (9.7 - 28.3), p<0.001 | 2.9 (2.5 - 3.3), p<0.001 | 4.7 (2.7 - 8.1), p<0.001 |
| IMD category Q2 (16.5 - 27.7) | 1.8 (1.4 - 2.2), p<0.001 | 2 (1.8 - 2.2), p<0.001 | ns |
| IMD category Q3 (27.8 - 39.0) | 1.7 (1.4 - 2.1), p<0.001 | 1.5 (1.4 - 1.7), p<0.001 | ns |
| IMD category Q4 (39.3 - 45.7) | 1.6 (1.3 - 2), p<0.001 | 1.4 (1.3 - 1.6), p<0.001 | ns |
| IMD category Q5 (45.7 - 71.8) | ns, p=0.517 | ns, p=0.574 | ns |
| Ethnicity South Asian | 0.4 (0.3 - 0.5), p<0.001 | 0.5 (0.4 - 0.5), p<0.001 | 1 (0.7 - 1.2), ns, 0.735 |
| Ethnicity Black | 1.7 (1.3 - 2.1), p<0.001 | 0.6 (0.5 - 0.7), p<0.001 | 3.1 (2.4 - 4), p<0.001 |
| Smoking Current or Ex | 0.3 (0.2 - 0.3), p<0.001 | 0.5 (0.4 - 0.5), p<0.001 | 0.7 (0.5 - 0.8), p<0.001 |
| Prior emergency admissions (1 year) | 1.6 (1.5 - 1.7), p<0.001 | 1.8 (1.7 - 1.8), p<0.001 | 0.9 (0.9 - 1), p<0.05 |
| Palliative care registered | 4.1 (3.2 - 5.1), p<0.001 | 1.7 (1.4 - 2), p<0.001 | 2.4 (1.9 - 3.1), p<0.001 |
| Nursing home resident | 1.7 (1.3 - 2.3), p<0.001 | 1.2 (1 - 1.5), ns, 0.058 | 1.7 (1.3 - 2.2), p<0.001 |
| Co-morbidities ≥3 | 1.5 (1.2 - 1.9), p<0.01 | 1.4 (1.3 - 1.6), p<0.001 | ns |
| Chronic obstructive pulmonary disease | 1.9 (1.5 - 2.3), p<0.001 | 1.8 (1.6 - 2), p<0.001 | ns |
| Peripheral vascular disease | 1.8 (1.4 - 2.3), p<0.001 | 1.2 (1 - 1.4), p<0.05 | 1.5 (1.1 - 2), p<0.01 |
| Atrial fibrillation | 1.7 (1.4 - 2.1), p<0.001 | 1.4 (1.2 - 1.5), p<0.001 | 1.3 (1.1 - 1.6), p<0.01 |
| Diabetes | 1.6 (1.4 - 1.9), p<0.001 | 1.3 (1.2 - 1.4), p<0.001 | 1.4 (1.2 - 1.6), p<0.001 |
| Cardiac failure | 1.6 (1.3 - 1.9), p<0.001 | 1.4 (1.2 - 1.6), p<0.001 | ns |
| Rheumatoid arthritis | 1.5 (1.1 - 2), p<0.01 | ns, p=0.134 | ns |
| Epilepsy | 1.4 (1 - 2), p<0.05 | 1.4 (1.2 - 1.6), p<0.001 | ns |
| Chronic kidney disease | 1.3 (1.1 - 1.6), p<0.01 | 1.2 (1 - 1.3), p<0.01 | ns |
| Hypertension | 1.2 (1 - 1.5), p<0.05 | ns, p=0.196 | 1.2 (1 - 1.4), p<0.05 |
| Cancer | 1.2 (1 - 1.5), p<0.05 | 1.6 (1.5 - 1.8), p<0.001 | 0.8 (0.7 - 1), p<0.05 |
| Cerebro-vascular disease | ns, p=0.101 | 1.2 (1 - 1.3), p<0.05 | ns |
| Ischaemic heart disease | ns, p=0.859 | 1.5 (1.3 - 1.6), p<0.001 | 0.7 (0.6 - 0.9), p<0.01 |
| Depression | ns, p=0.34 | 1.1 (1 - 1.2), p<0.01 | ns |
| Dementia | ns, p=0.059 | 0.8 (0.7 - 1), p<0.05 | ns |

**Figure 1:**
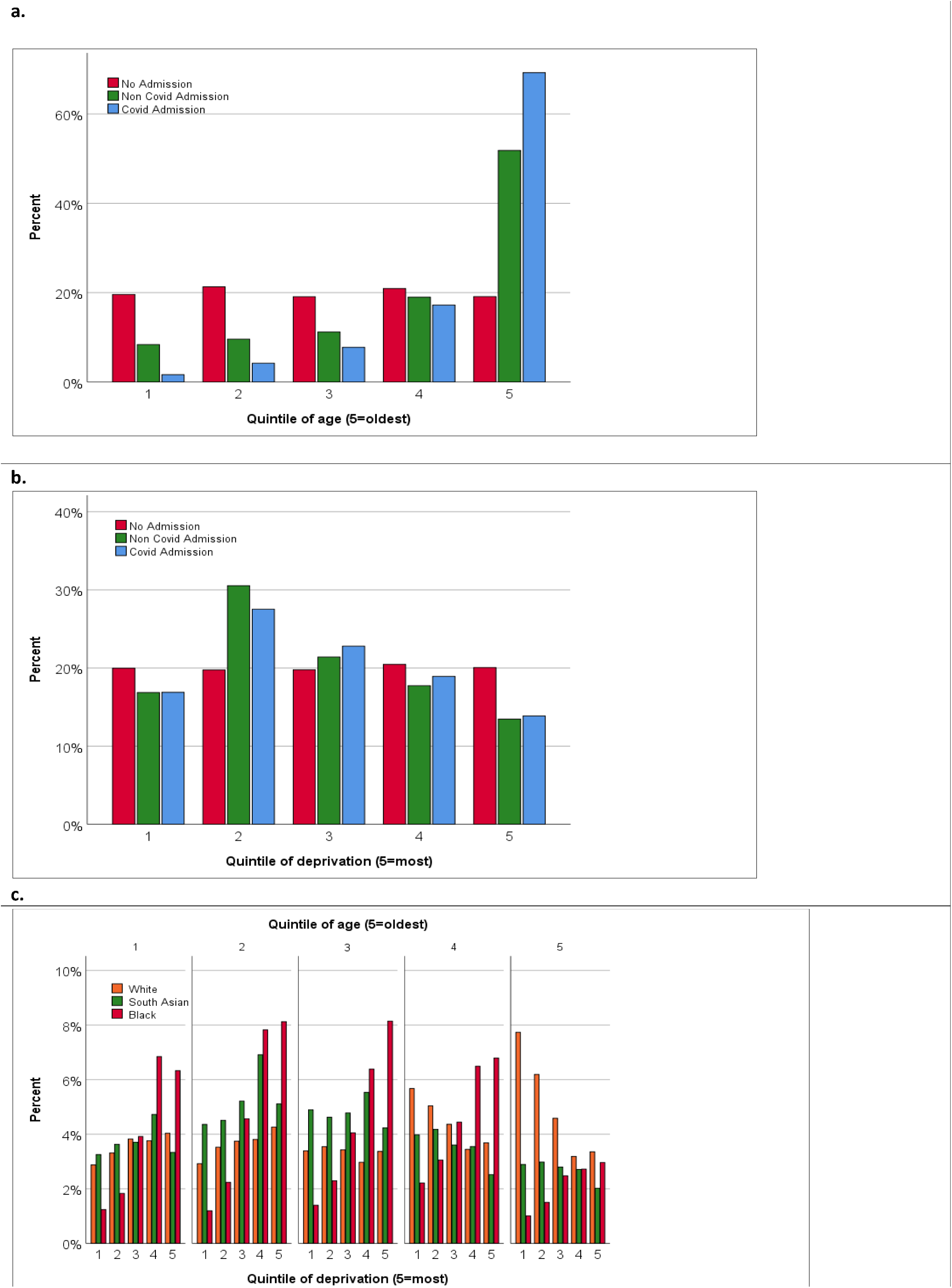
Age and deprivation in relation to hospital admission and in whole population. The association of age (**a**.) and deprivation (**b**.) with hospital admission type. Figure 1c. shows the inter-relationship of age, deprivation and ethnicity in the whole population (n=228,632) (Other / Unknown ethnic groups not shown).

The South Asian ethnic group was less likely to have a CA or NCA (60% and 50% crude percentage reduced risk respectively) compared to the White ethnic reference category whilst the Black ethnic group shared the significant propensity not to have an NCA but had a markedly increased relative risk (70%) for CA. Ethnicity related outcomes were examined specifically amongst those with COVID admission, by comparing those admitted to those not admitted within their ethnic category in separate binary regression analyses (all χ^2^ >252.4, p<0.001) (Table 4). Age, gender, preceding emergency admissions, palliative phase, comorbidity, and nursing home residence were significant associations in 2 or more of the ethnic groups. Of note, patterns of significantly associated individual comorbidities were different between the ethnic groups: Black - hypertension, atrial fibrillation and cardiac failure; South Asian - diabetes, peripheral vascular disease and atrial fibrillation; White - specific association with COPD, CKD, and RA. Deprivation had a significant impact only in the White group. The inter-relationship of age, deprivation with ethnicity and the impact of white ethnicity in the oldest quintile in lesser deprived categories can be seen in Figure 1c. In a simplified model of admission type and ethnicity (χ^2^=4542.9, p<0.001) with only age and deprivation entered categorically together with their interaction (χ^2^= 412.7, p<0.001), the ORs for CA compared to Whites were Black 2.08 (1.70 - 2.57) (p<0.001) and South Asian 0.56 (0.44 – 0.70) (p<0.001), with both groups still less likely to have NCA (p<0.001).

**Table 4.** Outcomes by individual ethnic category for those specifically with COVID hospital admissions compared those not admitted within individual ethnic grouping as examined in binary regression analysis. Values are the Odds Ratio with 95%CI.

| **Table 4** | **Black** | **South Asian** | **White** |
| --- | --- | --- | --- |
| Numbers admitted / not admitted | 113 (0.6%) / 17,530 | 83 (0.2%) / 43,663 | 684 (0.5%) / 138,500 |
| Gender (male) | ns | 2 (1.2 - 3.2), p<0.01 | 1.6 (1.4 - 1.9), p<0.001 |
| Age category Q2 (30 - 40) | 2.3 (0.6 - 8.6), ns, p = 0.225 | 1.8 (0.5 - 6.8), ns, p = 0.417 | 3 (1.3 - 6.8), p<0.01 |
| Age category Q3 (41 - 51) | 3.1 (0.9 - 11), ns, p = 0.083 | 2.6 (0.7 - 9.3), ns, p = 0.148 | 4.9 (2.2 - 10.5), p<0.001 |
| Age category Q4 (52 - 65) | 3.8 (1.1 - 13.4), p<0.05 | 3.1 (0.9 - 10.8), ns, p = 0.082 | 9.7 (4.7 - 20), p<0.001 |
| Age category Q5 (66 - 113) | 10 (2.8 - 36), p<0.001 | 4.5 (1.3 - 16), p<0.05 | 21.2 (10.4 - 43.3), p<0.001 |
| IMD category Q2 (16.5 - 27.7) | ns | ns | 1.7 (1.4 - 2.1), p<0.001 |
| IMD category Q3 (27.8 - 39.0) | ns | ns | 1.6 (1.3 - 2.1), p<0.001 |
| IMD category Q4 (39.3 - 45.7) | ns | ns | 1.5 (1.2 - 2), p<0.01 |
| IMD category Q5 (45.7 - 71.8) | ns | ns | ns |
| Smoking Current or Ex | 0.3 (0.2 - 0.6), p<0.001 | 0.2 (0.1 - 0.6), p<0.01 | 0.3 (0.2 - 0.3), p<0.001 |
| Prior emergency admissions (1 year) | ns | 1.5 (1.2 - 1.8), p<0.001 | 1.5 (1.4 - 1.6), p<0.001 |
| Palliative care registered | 3.8 (1.8 - 8), p<0.001 | 11 (5.5 - 21.7), p<0.001 | 3.8 (2.9 - 5), p<0.001 |
| Nursing home resident | ns | 4.5 (1.4 - 14.2), p<0.05 | 1.6 (1.1 - 2.2), p<0.01 |
| Co-morbidities ≥3 | 2.6 (1.5 - 4.4), p<0.001 | ns | 1.7 (1.3 - 2.1), p<0.001 |
| Atrial fibrillation | 2 (1 - 3.7), p<0.05 | 2.8 (1.4 - 5.6), p<0.01 | 1.6 (1.3 - 2), p<0.001 |
| Cardiac failure | 2.2 (1.2 - 3.9), p<0.05 | ns | 1.7 (1.3 - 2.1), p<0.001 |
| Chronic kidney disease | ns | ns | 1.4 (1.1 - 1.7), p<0.01 |
| Chronic obstructive pulmonary disease | ns | ns | 1.9 (1.5 - 2.3), p<0.001 |
| Diabetes | ns | 4.4 (2.6 - 7.5), p<0.001 | 1.5 (1.3 - 1.9), p<0.001 |
| Hypertension | 2.2 (1.2 - 4.1), p<0.01 | ns | ns |
| Peripheral vascular disease | ns | 3.7 (1.6 - 8.8), p<0.01 | 1.9 (1.4 - 2.5), p<0.001 |
| Rheumatoid arthritis | ns | ns | 1.6 (1.1 - 2.1), p<0.01 |

### Absolute risk of COVID Admission within Ethnic groups

The absolute risk from COVID hospital admission was 4.8 / 1000 population and Table 2 shows this broken down by ethnic grouping giving numbers, percentages, absolute risk and excess risk with unadjusted ORs compared to the White group, with the South Asian group showing a lower and the Black group a higher absolute risk as reflected in the unadjusted ORs.

### Mortality outcomes

COVID and non-COVID hospital death and death in the community (CHD, NCHD, DIC) were analysed in stepwise backwards multinomial regression (χ^2^=5,548.3, p<0.001) (Table 5). Male gender was significantly positively associated with mortality in all 3 categories. Increasing age was a significant factor, but there was no significant difference in age quintile distribution (χ^2^=12.168, p= 0.144, ns) with 89%, 84% and 86% in the oldest quintile in the CHD, NCHD and DIC groups respectively. For deprivation, for CHD and NCHD the pattern mirrored that of hospital admission with significantly increased mortality rates in the lesser deprived quintiles but not in the highest quintile whereas in DIC, a significant effect showing an increased mortality rate was only seen in the most deprived quintile. All categories shared a propensity for greater prior emergency admissions, multi morbidity and being in a palliative phase of care whilst being nursing home residency was associated with death in the community rather than hospital death. Individual morbidities varied in their associations, noting that diabetes and chronic kidney disease were in increased association with mortality only in the CHD group. The Black ethnic minority had significantly higher, and the South Asian significantly lower COVID hospital mortality rate, proportionately mirroring admission rates. Directly comparing CHD to NCHD confirmed a significantly increased association with Black ethnicity (OR 4.6 (2 - 10.2), p<0.003), diabetes (OR 1.5 (1 - 2.3), p<0.005) and chronic kidney disease (OR 1.6 (1.1 - 2.3), p<0.004) and an even greater negative association with current of previous smoking (OR 0.1 (0 - 0.3), p<0.002).

**Table 5.**
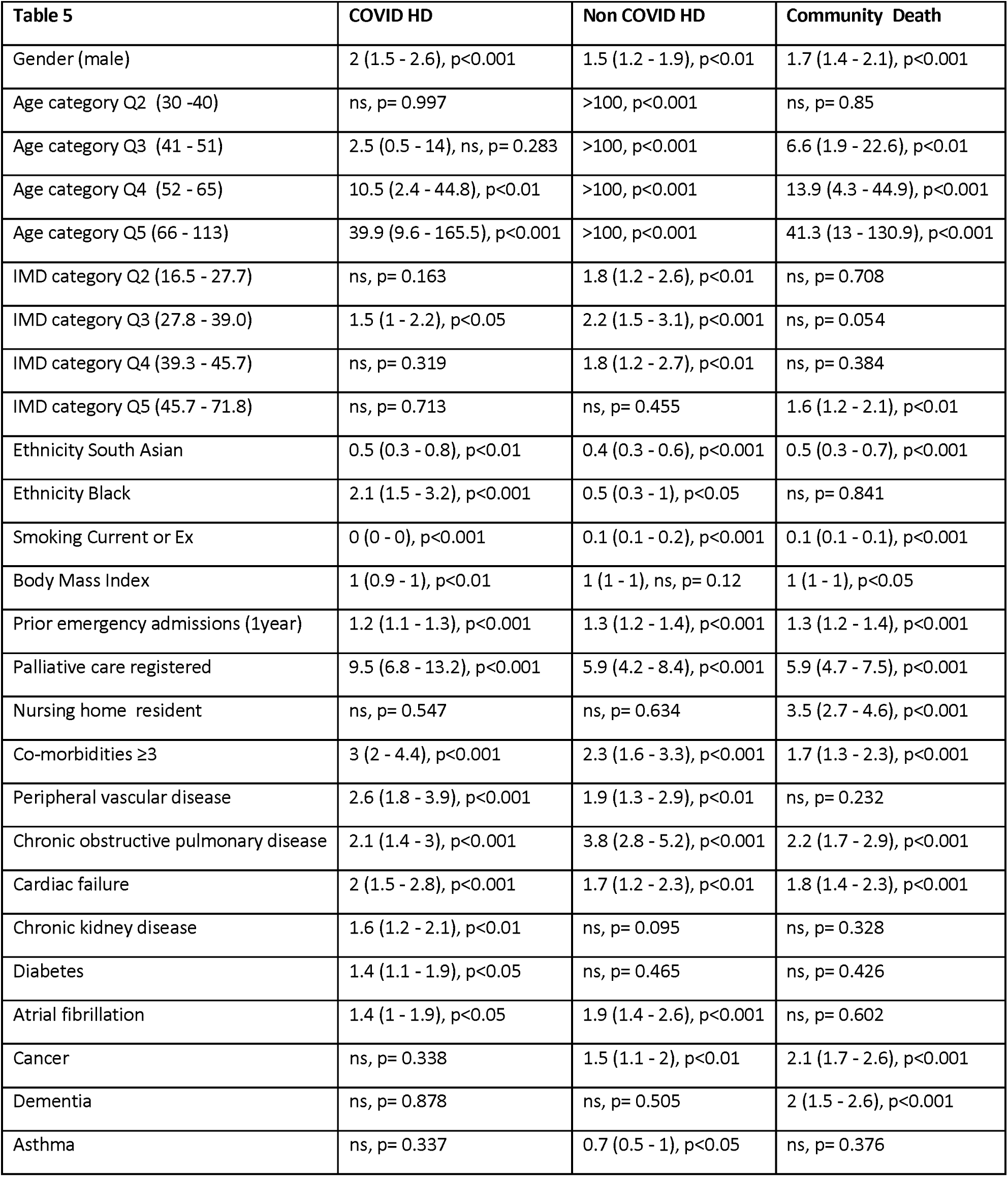
Multinomial regression of mortality outcomes over the 12 weeks period for those that died (n= 1093) either in the community (n= 537) or Non COVID (n = 284) and COVID related (n= 272) hospital deaths (HD) (See Table 1) compared to those who were alive (n = 227, 539). Results are the Odds ratio with 95% confidence intervals. For age and IMD categories the comparators were the youngest and least deprived quintile respectively. Variables not listed from Table 1 were excluded stepwise (backwards) as not significant.

### Absolute risk of COVID Death by Ethnic group

Specifically for COVID death, Table 2 shows numbers, percentages, absolute risk and excess risk with unadjusted ORs for the ethnic minorities compared to the White group and Figure 2a shows the distribution of mortality outcome by ethnic category (χ^2^ = 126.1, p<0.001). The absolute risk of COVID death was 1.32, 0.73 and 2.2 per 1000 population in the White, South Asian and Black ethnic groups and the excess risk was -0.61 (negative) and 0.85 deaths per 1000 population in South Asians and Blacks versus Whites respectively. Compared to the White population, the unadjusted OR (95% CI) for COVID death for the Black and Asian groups was 1.6 (1.2 – 2.3) and 0.5 (0.4 – 0.8) respectively (both p<0.01). The ethnic groups differed significantly in age (White 50±20, South Asian 45±16, Black 45±17 years, F=1868.9, P<0.001) and age was the dominant factor associated with hospital admission and death (Tables 1, 3, 4, 5). To avoid any potential misrepresentation of mortality outcomes by statistical age adjustment, the absolute effects were considered for the oldest quintile only where 84% of all COVID deaths occurred, in which case the ORs were Black 3.9 (2.7 – 5.6) (p<0.001) and South Asian 0.9 (0.6 – 1.4) (p=0.72, ns) (Figure 2b).

**Figure 2:**
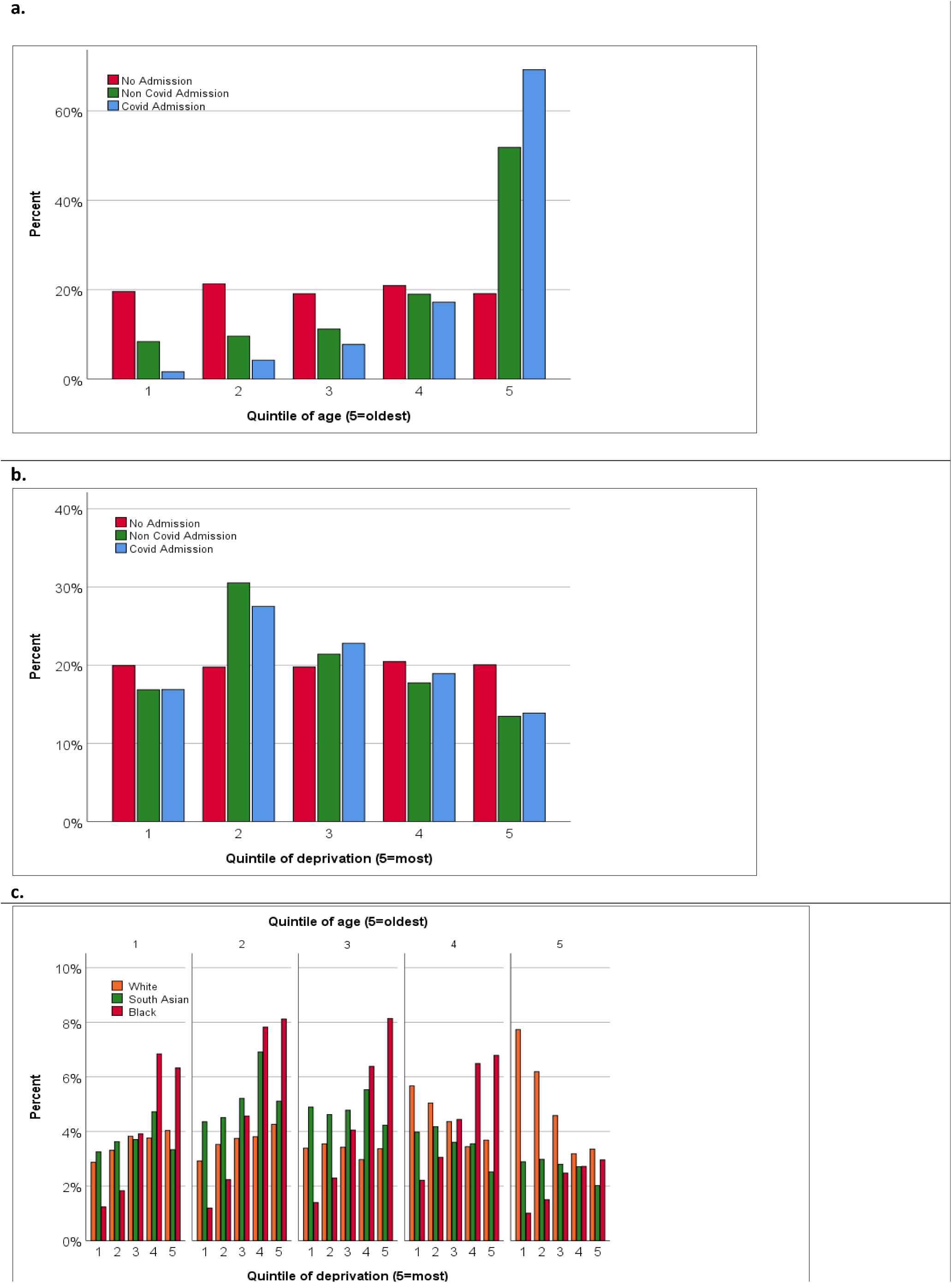
Mortality by ethnicity. Crude mortality by ethnic grouping as percentages (**a**. χ^2^ = 184·4, p<0·001), within the oldest quintile (**b**. χ^2^=92·2, p<0·001) or restricted to those with a COVID admission excluding those with a non-COVID death (**c**. χ^2^=5.92, p=0.115, ns), (Other / Unknown ethnic categories are not shown but were included in the analysis).

### COVID Hospital admission and COVID mortality

By introducing hospital admission status, the COVID mortality ORs were Black 1.3 (0.9 – 2.0) (p=0.206, ns) and South Asian 1.5 (0.9 – 2.3) (p=0.098, ns) were similar, indicating similar in hospital mortality in contrast to the whole population effect. To negate this potential effect of prior propensity for acquisition of serious COVID infection, and focusing on the Black and South Asian minorities compared to the White majority, a narrower assessment of those who were admitted with COVID and had a COVID death was made. Amongst 930 COVID admissions, excluding those with a COVID admission but with non-COVID death (n = 83 (9%), COVID death occurred 270 (32%) (White 189, South Asian 32, Black 38, Other 11). The ORs for the association of ethnicity with COVID mortality were Black 1.2 (0.7 – 1.8) (p=0.423, ns) and South Asian 1.6 (1.0 – 2.5) (p= 0.075, ns) (χ^2^= 5.92, p= 0.115, ns) (Figure 2c). Utilising the full model with all independent variables, including age, which remained significantly different between ethnic groups (F= 13.23, p<0.001), then the significantly associated variables were age, gender, smoking status, body mass index, palliative phase of life, multi-morbidity and the individual comorbidities of cardiac failure, chronic kidney disease and peripheral vascular disease but not ethnic grouping or deprivation score (Table 6). Finally, Table 2 shows the absolute risks of COVID death in COVID hospital admission and unadjusted ORs which are consistent with the findings of the modelled data.

**Table 6.** Multinomial regression amongst those with a COVID admission restricted to the White, Black and South Asian ethnic groups (n=797) comparing to those with COVID death (n= 259) to those who were alive at 12 weeks. Results are the Odds Ratio with 95% confidence intervals. Variables not listed from Table 1 were excluded stepwise (backwards) as not significant.

| **Table 6** | **OR COVID death vs alive** |
| --- | --- |
| Age | 1.05 (1.03 - 1.07), p<0.001 |
| Gender (male) | 1.91 (1.3 - 2.83), p<0.01 |
| Smoking Current or Ex | 0.01 (0 - 0.04), p<0.001 |
| Body Mass Index (kg/m2) | 0.92 (0.86 - 0.98), p<0.01 |
| Palliative care registered | 7.83 (4.21 - 14.56), p<0.001 |
| Co-morbidities ≥3 | 1.64 (1 - 2.67), p<0.05 |
| Cardiac failure | 1.82 (1.14 - 2.92), p<0.05 |
| Chronic kidney disease | 1.69 (1.09 - 2.63), p<0.05 |
| Peripheral vascular disease | 2.27 (1.12 - 4.59), p<0.05 |

## Discussion

### Principal findings

Over and above known general associations with hospital admission and mortality, our study suggest a complex association of deprivation and points to heterogeneity of the impact of ethnicity, both of which may vary by locality. We highlight the need for local health economies to have robust, accurate and integrated clinical data in order to assess and inform local decisions making and, in particular, at a time of heightened anxiety, we raises a concern about the conveyance of risk to local communities. The crucial differences in relationship to other studies are as follows:

### General Associations

Uncontroversially, factors associated with non-COVID or COVID hospital admission and death included age, gender, prior emergency admissions, and palliative phase of life, nursing home residence and multi-morbidity with specific comorbidities associated with COVID admission or death and with ethnic status. Any association of smoking with better COVID outcomes, observed in other studies, ^2,17^ may be refuted when taken in the context of this being common to non-COVID admissions and death during this period.

### COVID vs Non COVID Admissions

The significant differences were age, gender and degree of comorbidity complexity (palliative care, nursing home) but as it is likely that patterns of emergency admissions differed at this time, comparisons of COVID to non-COVID hospital admission may have little relevance to COVID outcomes, noteworthy for studies that have reported on COVID hospital admission alone.^12,13,18^

### Deprivation

For hospital non-COVID and COVID admission and death, the pattern was for excess in lesser deprived quintiles in the White ethnic population but not within ethnic minority groups where deprivation was not a significant factor. This contrasts with other studies: ^2, 5^ in some deprivation was not a significantly associated factor in fully adjusted models,^19^ whilst other UK studies, ^18^ and most overseas studies, have not considered this.^12, 13^ Following the H1N1 pandemic influenza of 2009, many studies indicated effects of deprivation including a rural urban divide impact,^20^ as is seen in this pandemic.^21^ Our findings within a health economy with significant deprivation call for the need to explore this association within larger studies specifically within urban areas.

### Ethnicity

We note that a recent meta-analysis shows heterogeneity in the association of ethnicity to COVID mortality;^22^ in a large population study reporting adverse odds ratios for all ethnic groups, their crude unadjusted data showed significantly increased risk in the Black (χ^2^ = 17.464, p<0.001) but not in the South Asian group (χ^2^ =3.238, p=0.072),^2^ as shown in another population level study;^21^ in the largest reported hospital admission series both ethnic groups had significantly lower unadjusted mortality rates,^15^ whilst in their modelled data no effect was seen amongst Blacks; from New York there was no adverse ethnicity signal,^12,13^ and early reported adverse ethnicity outcomes in the 2009 UK flu pandemic,^23^ did not stand subsequent review.^24^ We find our Black population had significantly higher and the South Asian lower crude and adjusted COVID admission rates compared to Whites, also observing that both ethnic subgroups had lower non-COVID hospital admissions, further contextualising the strong effect in the Black cohort. In both groups, their crude and adjusted patterns of COVID mortality mirrored that of COVID hospital admission but from the numerical base of COVID admission, there was no significant difference between the Black and South Asian compared to the White groups, highlighting pitfalls of examining effects in isolation. Our data in the Black population is broadly in keeping with some studies showing excess COVID hospitalisation and mortality but the South Asian group’s lower absolute and adjusted rate of admission and death from COVID 19 are strikingly different. Given the variation in findings to date, we do not consider this an “unexpected finding”, and hypothesise that many local population factors are at play including population density, family size, housing, duration of immigration, country of birth (including UK born) and occupation and the precise ethnic group within the ‘South Asian’ population may well be of importance. A recent updated analysis by the UK Office for National Statistics has emphasised that “*ethnic differences in mortality involving COVID-19 are most strongly associated with demographic and socio-economic factors, such as place of residence and occupational exposures, and cannot be explained by pre-existing health conditions*” which conclusion is consistent with our locally-dictated findings.^25^ Otherwise within BAME groups we find specific individual comorbidities vary in their association with COVID risks: in South Asians these are diabetes, peripheral vascular disease and atrial fibrillation; in the Black population hypertension, atrial fibrillation and cardiac failure; in white ethnicity most co-morbidities but in particular COPD, chronic kidney disease and rheumatoid arthritis.

### The conveyance of risk

Public health messages are vital to convey but population adjusted risk rates may confuse, adversely impacting behaviors such that, it is feared, hospital admission patterns may change unfavourably. Absolute, absolute excess, relative, unadjusted and adjusted risk is complex to communicate even for healthcare professionals making them susceptible to reasoning errors and misinterpretation of probabilities ^26^ and individuals, with erstwhile health risk, should know about the magnitude of risk in a way that can be conceptualised.^27,28^ For our Black population, the fully-modelled OR for COVID mortality was 2.1, the absolute risk 2.2 / 1000 people or an excess risk of 0.9/1000. A Black person in Wolverhampton ought to be informed that “twice as likely to die of COVID” compared to the White community can also mean “a 1 in 1000 excess risk”.

### Strengths and Weaknesses

Combining Wolverhampton’s health data evaluated our local population’s heterogeneous demographic and its associations with community or hospital non-COVID and COVID hospital admission and mortality, uniquely approaching these outcomes simultaneously. This local nuance complements larger studies, informing appraisal of risk from an urban, and multi-ethnic and deprived setting, highlighting concerns of extrapolation from larger datasets to UK localities. An example of a particular strength of the data quality was the cross check ascertainment of COVID admission, without sole reliance on COVID testing, permitting specific categorisation of deaths (COVID, non-COVID and post discharge) rather than less accurately into global mortality.

Limitations of the study: This is a twelve-week evaluation spanning the pandemic’s upsurge and peak; the population and event number were comparatively small; cause of death in the community was unknown and it is likely that people died away from the hospital undiagnosed with COVID 19 ; some data were missing but this was mitigated; whilst being aligned to the population at 99.5% concordance, hospital data were not totally drawn from the City population, which varied by GP registration, residency, or admission from immediately surrounding areas and a small proportion of admissions were non-resident or non-registered, so this is not strictly an epidemiological study but an observational study comparing defined cohorts in tiers of analysis (e.g. COVID death amongst COVID admissions) where this caveat does not apply.^16^

### Implications for clinicians and policy makers

We show that a variety of recognised factors were associated with COVID death, as with non-COVID death. At our local level, COVID admission and death were not strongly associated with worsening deprivation, with a novel potential different relationship in the White population.

Higher absolute and adjusted COVID admission and mortality occurred in the Black population whilst they were reduced in Wolverhampton’s South Asian community. We point out the non-significant association between in-hospital COVID-19 case fatality and ethnicity, raising the probability that COVID-19 mortality relates to differential risks of exposure, susceptibility and disease contraction before hospital admission, let alone the possible avoidance of hospital admission. Two important considerations are the potential excessive use of multiple factors and the disruption of the perspective from a population’s base through hospital admissions to COVID specific admission, leading to widely varying conclusions, highlighting the difficulties of using observational data and the potential for Collider bias.^29^ We support the case for more localised population-based studies of both hospital admission and subsequent death, such as ours, in which the denominator and numerator populations can be clearly linked and are fully and transparently ascertained and characterised. To avoid associations in the data being due to the way in which data are sampled, local health economies should be mandated to link hospital and primary care data across their population level down to the un-anonymised individual level; they should, in preparation for future epidemics, have data quality mechanisms in place to ensure accuracy in their demographics, the accrual of important missing data and the triangulation of key outcomes to minimise false positive and negative results. This includes the need to have a robust, systematic, accurate and timely approach to the recording of death whether in the community or hospital setting. A defined data set and its capture in routine clinical systems seems apposite.^30^ Accepting that variation in findings in different population subsets is both inevitable and valid, we would suggest the need for the public health and research community to accommodate uncertainty in emergent evidence, learning from the experience of previous viral pandemics. This includes the need to have a robust, systematic, accurate and timely approach to the recording of death whether in the community or hospital setting.^31^

### Future research

Perhaps most crucially we argue that, in reporting future research of this kind during the current pandemic and beyond, there is an ethical obligation for the standardisation of the conveyance of risk in a manner that spans the absolute to the relative so that is easily comprehensible to the individuals and populations at risk and others, including health professionals, politicians and the media, all matters in which the editorial and peer review mechanism of our medical journals have a vital role.

## Data Availability

Anonymised data may be obtained on reasonable request from the corresponding author

## Author contributions

Accountable senior author BMS; Data analysis, manuscript writing: BMS, JB, SJD, AV; Preparation for submission: SJD, BMS, JB; Database quality, data integration and data quality and integration: AV, BMS, VK; Reading drafts as lay expert: SM; Statistical advice: AN; All authors contributed intellectual content during the drafting and revision of the work and approved the final version.

## Transparency declaration

BMS affirms that the manuscript is an honest, accurate, and transparent account of the study being reported; that no important aspects of the study have been omitted; and that any discrepancies from the study as planned (and, if relevant, registered) have been explained

## Data Sharing

Anonymised data will be shared on reasonable request to the corresponding author

